# 68 Consecutive patients assessed for COVID-19 infection; experience from a UK regional infectious disease unit

**DOI:** 10.1101/2020.02.29.20029462

**Authors:** Nicholas Easom, Peter Moss, Gavin Barlow, Anda Samson, Thomas Taynton, Kate Adams, Monica Ivan, Phillipa Burns, Kavitha Gajee, Kirstine Eastick, Patrick J Lillie

**Author notes:** Corresponding author Dr Patrick Lillie, Department of Infection, Hull University Teaching Hospitals NHS Trust, Castle Hill Hospital, East Yorkshire, HU16 5JQ, United Kingdom, Email –.

## Abstract

Clinical assessment of possible infection with SARS-CoV-2, the novel coronavirus responsible for the outbreak of COVID-19 respiratory illness, has been a major activity of infectious diseases services in the UK and elsewhere since the first report of cases in December 2019. We report our case series of 68 patients, reviewed by Infectious Diseases Consultants at a Regional Infectious Diseases Unit in the UK. We prospectively evaluated our service between the 29^th^ Jan 2020 and 24^th^ Feb 2020.

Demographic, clinical, epidemiological and laboratory data were collected. We have compared clinical features and subsequent diagnosis between well patients not requiring admission for clinical reasons or antimicrobials with those assessed as needing either admission or antimicrobial treatment.

Final microbiological diagnoses included SARS-CoV-2 (COVID-19), Mycoplasma pneumonia, influenza A, RSV, non SARS/MERS coronaviruses, rhinovirus/enterovirus. 9/68 were treated with antimicrobials, 15/68 were admitted to a negative pressure room of whom 5/68 were admitted solely due to an inability to isolate at home. Patients requiring either admission on clinical grounds or antimicrobials (14/68) were similar to those not requiring admission or antimicrobials, with modestly more fever and shortness of breath in the clinically admitted / antimicrobial group. The most commonly prescribed antimicrobials were doxycycline, moxifloxacin and oseltamivir.

The majority of patients had mild illness which did not require a clinical intervention to manage. This finding supports a community testing approach supported by clinicians to review the proportion of more unwell patients.

## Introduction

SARS-CoV-2 is a recently named novel coronavirus responsible for the outbreak of respiratory disease named COVID-19, arising in Wuhan, China, in December 2019^1-3^. At the time of writing (27^th^ Feb 2020) the overwhelming majority of cases have been reported from inside China, however single or small numbers of cases have been confirmed in 46 other countries. Transmission has occurred in some countries outside China^4,5^, including the UK^6^.

In the UK, public health and clinical services have been working to identify suspected cases according to a national case definition and to arrange testing, predominantly by real-time PCR of nose and throat swabs. Since testing began, local procedures and national guidelines have changed in response to changing understanding of the disease and demand for testing. On the evening of 06/02/2020 the UK definition of a suspected case was extended to include people presenting with respiratory illness (defined as cough, shortness of breath or fever with or without other symptoms) returning from or transiting through China including Hong Kong and Macau, Japan, Malaysia, South Korea, Singapore, Taiwan or Thailand within the last 14 days, with the case definition subsequently changing further on 25/02/2020 to include northern Italy, Iran and further countries in SE Asia. As of 27/02/2020, 7690 tests have been performed nationally, of which 15 were positive (0.2%). Initially, testing has been led by clinicians, predominantly by infectious diseases or emergency department physicians, although there are plans to move to a community testing model led by other groups of healthcare professionals, and some regions have already done so. Patients are instructed to self-isolate while results are pending and until their symptoms have resolved, with possible financial and health implications for the 99.8% so far in the UK who have an illness other than COVID-19. During the 2009 H1N1 influenza pandemic when a syndromic management strategy with presumptive treatment and self-isolation was used, initial clinical diagnoses of influenza were reported to delay diagnoses of a number of diseases including primary HIV infection^7^ and *Plasmodium falciparum* malaria^8^, and although scoring systems were developed it remains difficult to distinguish between viral and bacterial pneumonia on clinical grounds^9^. In addition, many mild respiratory viral infections were managed as influenza^10^, with significant resource implications, both for healthcare services and patients

Here we describe our experience of the first 68 patients we have tested for SARS-CoV-2 at a Regional Infectious Diseases unit (RIDU) in the UK. We present the spectrum of illness, alternative diagnoses made and management provided. This is of particular interest at this stage of the epidemic, where many individuals meeting the definition of a suspected case are returning travellers, where the differential diagnosis of respiratory or undifferentiated febrile illness may be broad^11^. These findings have implications for the clinical and logistical support that may be required for roll-out of community testing to be a safe and effective replacement for the current predominantly hospital-based, physician-led system.

## Methods

### Patients

The RIDU at Hull University Teaching Hospitals NHS Trust is based at Castle Hill Hospital, East Yorkshire, UK and serves a population of 1.2 million people. Patients were predominately referred following telephone assessment by the national NHS 111 service, using Public Health England (PHE) case definitions. Patients were assessed by an infection clinician either in the infectious diseases ward, the ambulance or the patient’s car that had transferred them to the unit, and in one instance in the emergency department of the trust. The first 68 consecutive cases are presented here.

### Clinical assessment and testing

All patients had symptoms recorded, together with a travel and exposure history for at least the 14 days preceding symptom onset. Due to the use of personal protective equipment and in some cases assessment in the patient’s car, clinical examination and observations varied between cases. Patients were managed as outpatients if clinically stable and able to self-isolate. All cases were tested for SARS2-CoV using a combined throat and naso-pharyngeal (NP) swab, which were processed at the designated public health laboratory. A separate NP swab was tested locally using the BioFire Film Array Respiratory Panel 2 plus (BioMérieux, Marcy l’Etoile, France) which can detect 21 targets (17 viral and 4 bacterial). Blood samples were only performed on patients being admitted or where another serious diagnosis was being considered.

### Data collection, analysis and governance approval

Data were entered directly from clinical notes into a centrally held password protected spreadsheet, to facilitate patient follow up and data analysis. Clinical features, observations, investigations, management and outcomes are presented with descriptive statistics where relevant. Local clinical governance approval was granted to record the data as an ongoing service evaluation. As all care delivered to patients was routine, no ethical approval is necessary in keeping with UK national guidance that this is an evaluation of a current NHS service.

## Results

### Timeline of cases, countries visited and location of management

The timeline of cases seen is shown in figure 1. With the change in case definition to include those returning from SE Asia, the number of cases markedly increased from an initial mean of 0.9 cases / day to 3.3 / day. 15 / 68 patients (23%) were admitted to the unit, with the remaining patients being managed in an ambulatory manner. Of those admitted, six were only admitted as they could not effectively self-isolate pending results. Mean length of hospital stay in those admitted was 2.4 days (range 1 – 10 days). 10 patients (15%) had returned from China in the 14 days prior to onset of symptoms, with 26 of the cases (38%) having visited the most common country of travel, Thailand. Eight patients had not left the UK in the preceding 14 days and were seen as contacts of confirmed or suspected cases.

**Figure 1.**
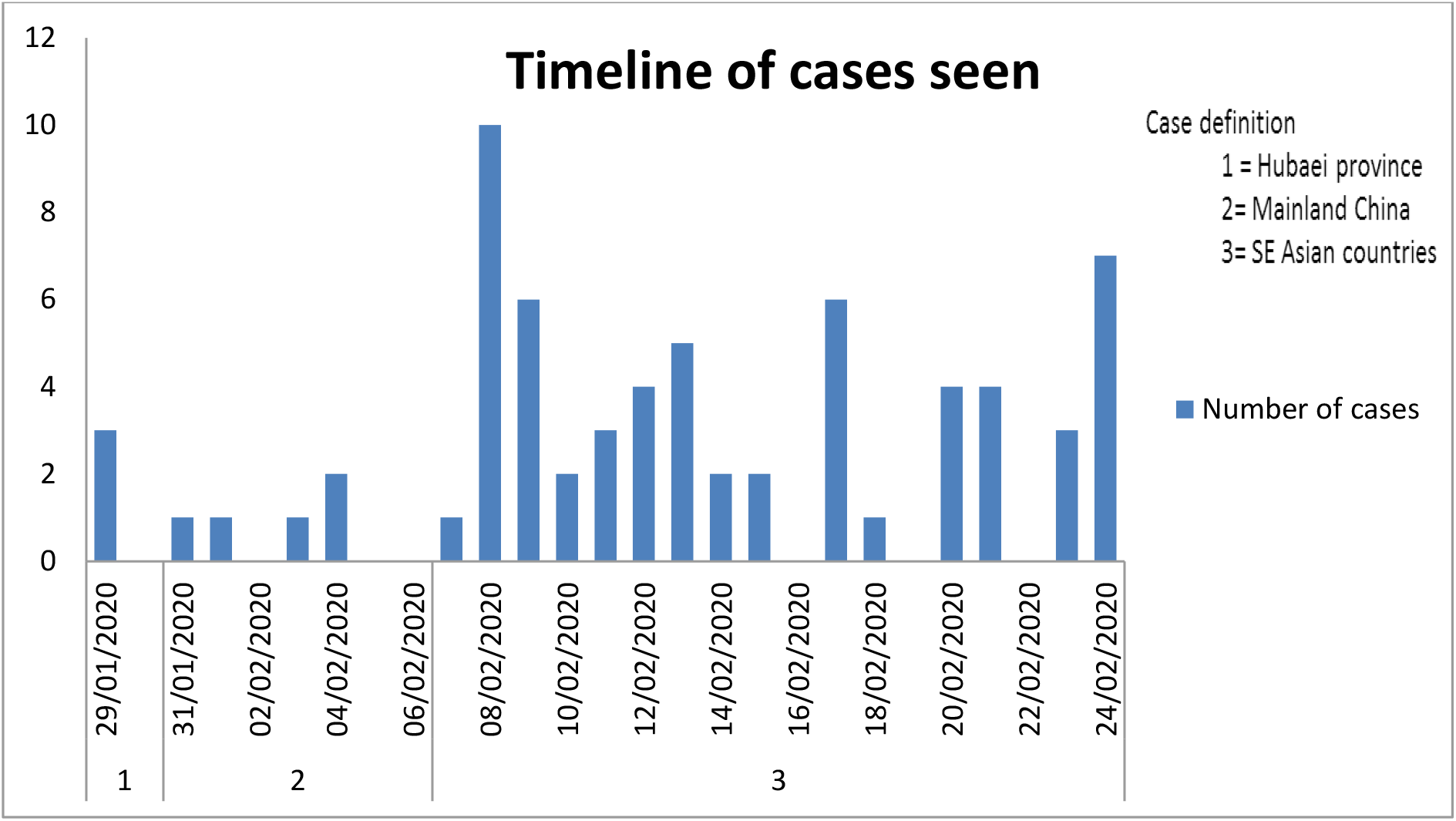
Timeline of cases

### Clinical features, patient demographics, antimicrobial usage / management and discharge diagnoses

Of the 68 patients seen 36 were female (53%), with a mean age of 42.5 years (range 0.5 – 76). Table 1 shows the presenting features of the cases seen. In addition to the symptoms shown in the table, three cases (4%) had headache, three (4%) had ear pain and nine (13%) had diarrhoea (two with vomiting as well). Table 2 shows the baseline physical observations, with seven patients (10%) having a temperature of 37.5° or greater on assessment. Antimicrobial therapy was prescribed to nine patients (1.3%), with doxycycline given to five patients (78%), moxifloxacin to three (4%), and oseltamivir to one confirmed influenza A case (1%).

**Table 1.**
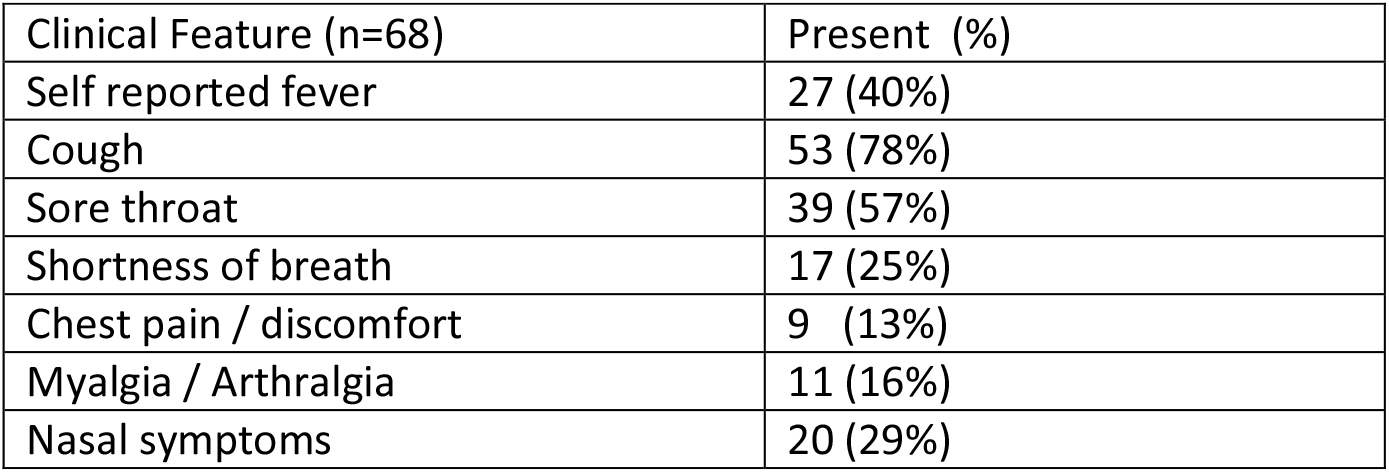
Presenting features of cases

| Clinical Feature (n=68) | Present (%) |
| --- | --- |
| Self reported fever | 27 (40%) |
| Cough | 53 (78%) |
| Sore throat | 39 (57%) |
| Shortness of breath | 17 (25%) |
| Chest pain / discomfort | 9 (13%) |
| Myalgia / Arthralgia | 11 (16%) |
| Nasal symptoms | 20 (29%) |

**Table 2.** Observations of cases (excluding 2 children assessed)

| Observation | Mean | Range |
| --- | --- | --- |
| Temperature (°C) (n=58) | 36.4 | 33.5 – 38.8 |
| Respiratory rate (/ minute) (n=57) | 17 | 14 – 24 |
| Oxygen saturations on air (%) (n=56) | 97.6 | 94 – 100 |
| Pulse (/minute) (n=58) | 83.2 | 52 – 125 |
| Systolic blood pressure (mm Hg) (n=45) | 125 | 96 – 180 |

Clinical diagnoses in this cohort included upper respiratory tract infection in 50 patients (74%) exacerbations of airways disease in five patients (7%), lower respiratory tract infections in four patients (6%), gastroenteritis in three (4%), influenza like illness in two patients (3%), with one patient (1%) each diagnosed with the following: otitis media, a well contact, inebriation and community-acquired pneumonia.

### Laboratory investigations

Table 3 shows the results of both virological sampling for standard respiratory pathogens in 49 (98%) of the cases assessed. In addition to the virological sampling of NP swabs, four patients (6%) had sputum tested using the BioFire pneumonia panel, with one patient (2%) found to have *Staphylococcus aureu*s (MSSA), one patient with *E*.*coli* and *Haemophilus influenzae*, one patient with *Haemophilus influenzae* and seasonal Coronavirus co-infection, and one with *Haemophilus influenzae* mono-infection. Malaria films were performed and were negative in four patients (6%), whilst blood cultures were negative in all 14 patients (28%) they were performed on. Standard clinical chemistry and haematology tests were performed in 20 patients (29%). Five patients (7%) had elevated total white cell counts (highest recorded 15.3 × 10^9^ / L) with one patient having mild thrombocytopaenia (92 × 10^9^ / L). No patients had abnormal serum creatinine or electrolytes, with two (4%) having mildly elevated serum alanine transaminase levels (highest level 63 IU / L). C-Reactive Protein (CRP) levels were within the normal range in 12 out of 19 cases tested (63%), with the highest recorded level being 73 mg / L.

**Table 3.**
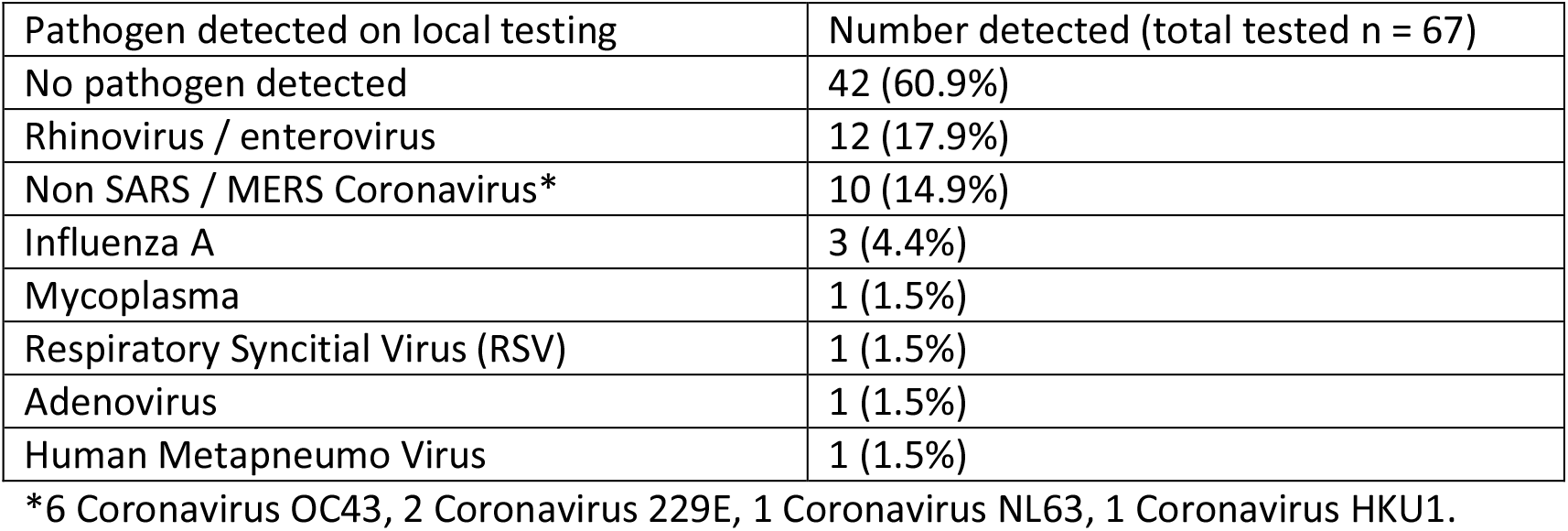
Results of multiplex PCR of nasopharyngeal swabs

### Comparison of clinically well patients with those requiring antimicrobials / clinical need for admission

Table 4 shows the physiological and demographic features of those patients who were prescribed antimicrobial therapy OR admitted for clinical reasons (as a group that represents those patients requiring medical input) compared to the other patients seen who were either managed as outpatients, or admitted due to being unable to self-isolate. Fever ≥37.5° and shortness of breath as a symptom were the only predictors of requiring antimicrobials or admission for a clinical need.

**Table 4.** Differences between patients requiring medical admission OR antimicrobials

|  | Antimicrobial / medical admission group (n= 14) | Remaining patients (n= 52) | P value (Fishers exact test / t-test) |
| --- | --- | --- | --- |
| Age (yrs) | 42.2 | 42.6 | 0.949 |
| Temperature $\geq 37.5^{\circ}$ | 5 / 14 | 2 / 46 | 0.0057 |
| Pulse (/min) | 88.6 | 81.8 | 0.225 |
| Respiratory rate | 17.9 | 16.7 | 0.0316 |
| Reported shortness of breath | 6 / 14 | 11 / 54 | 0.0968 |
| Cough | 9 / 14 | 44 / 54 | 0.2753 |

(Physiological parameters did not include the two children assessed)

## Discussion

We have presented our experience of testing for SARS-CoV-2 at a UK university teaching hospital. Sixty-eight individuals, including 2 children, were tested over 26 days. The majority were ambulant and able to self-isolate, and had features consistent with an upper respiratory tract viral infection, or common cold. After the first 10 cases, 6/58 (10.3%) were admitted, at least three of whom only because they lived in shared accommodation where self-isolation was unrealistic, rather than on clinical grounds. This observation supports a community testing approach for the majority. Five of fifty individuals were prescribed antimicrobials without being admitted, emphasising the importance of the availability of clinical decision makers and prescribers for more unwell patients. None of our cases had severe respiratory illness requiring non-invasive or mechanical ventilation, suggesting it will be rare that patients meeting the definition of a suspected case will require enhanced care, at least while COVID-19 itself remains rare in the UK. Some COVID-19 case series have suggested up to 20% of cases may require respiratory support^2,12^, although this figure may be influenced by ascertainment bias. If more generalised outbreaks occur outside China, we may start to see severe COVID-19 presenting to UK healthcare services.

Specialist Infectious Diseases consultant-delivered assessment of a group of patients who predominantly have mild illness is unlikely to be sustainable, especially as the case-definition broadens to include a wider geographical area and/or COVID-19 patients requiring inpatient care becomes more common in the UK. There was a step-change in the number of suspected cases seen after the introduction of the third case definition on the evening of 6^th^ February 2020 as a high number of people with respiratory symptoms and recent travel suddenly became suspected of having COVID-19. As there is further spread in the coming weeks, as is the case in Italy and Iran, and the epidemiological criteria of the case definition is notably extended again, Infectious Diseases clinicians will be unable to sustain a hospital-based screening service. The NHS is responding to this challenge by moving to a community provider screening model, which will deliver the majority of testing in the home.

Specialist input and assessment, supported by appropriate laboratory investigations must be easily accessible to a COVID-19 testing service, especially if delivered by health care workers other than Infectious Diseases physicians. This is important for patients in the self-isolation periods both before and after receiving a test result as these individuals will not readily be able to access the usual healthcare services. This may apply particularly to patients with pre-existing lung disease, where respiratory viral illness, caused by SARS-CoV-2 or otherwise, may trigger clinical deterioration that could ordinarily be managed by early community intervention. Infectious Diseases physicians may be the group best placed to support patients in this situation for the short period of self-isolation,^13^ especially given the need for ongoing personal protective equipment use^14^. Although we did not observe any imported “tropical” infections such as dengue, malaria, typhoid, rickettsiosis or leptospirosis, these remain important differential diagnoses in returning travellers from some areas of South East Asia and may be life-threatening if missed^8,15^. These diagnoses may become more common in this group now that the definition of a suspected case has been expanded to include other countries in South or South East Asia such as Vietnam. Individuals receiving testing for COVID-19 should be signposted to an accessible clinical service with an understanding of this differential diagnosis in case of worsening illness.

Our data from the routine virological testing performed is in keeping with community based results showing that symptoms and viral detection do not always go together^16^ and shows a different spectrum to those patients admitted and tested using the same assay^17^, with fewer influenza diagnoses and a greater proportion of rhinovirus / enterovirus as well as seasonal coronaviruses cases. A recent report from Italy^18^, immediately before the recent increase in SARS-CoV-2 detection showed a greater proportion of influenza B than in our cohort, which may be explainable by the differing geographical exposures between the two groups.

Admitting individuals to a secondary care facility, even for short periods of assessment, may have other drawbacks. Travellers to South East Asia, particularly those who received antibiotics or were admitted to healthcare facilities, can potentially carry and transmit carbapenem-producing enterobacteriaceae. Additionally, because these patients need to be more pragmatically managed than patients not suspected of COVID-19, they may be more likely to be exposed to antimicrobial therapy. There are also significant resource implications not just in terms of PPE use but predominantly in terms of the time required to don and doff PPE for every care interaction from bringing food to routine observations to clinical review. This burden can be at least partially alleviated through a home testing approach.

Infection clinician time is another important resource that may not be best used in managing mild respiratory illness that meets the suspected COVID-19 case definition. In recent years, many studies have demonstrated the benefit of ID physician review in terms of clinical outcome and resource utilisation in infections such as *Staphylococcus aureus* bacteraemia^19,20^ and resistant gram-negative infections^21^. Our findings support the assessment of, and testing for, possible COVID-19 by other cadres of healthcare workers, outside the secondary care setting, but supported by specialist physicians and hospital based further diagnostics where needed. The optimum configuration for such a service remains unclear and is likely to vary depending on local constraints, but during the containment phase of the epidemic response there may be time to pilot and compare a range of models before we are forced to move to delay and possibly mitigation strategies.

## Data Availability

Data available inf needed with identifiable data redacted (current spreadsheet in use for patient care as well as data collection)

